# Compatibility and Bias Between Ultrasound-Guided Attenuation Parameter and Controlled Attenuation Parameter for Ultrasound-Based Hepatic Steatosis Assessment

**DOI:** 10.1101/2025.06.03.25328166

**Authors:** Daisuke Miura

## Abstract

**Purpose:** To develop a regression model for estimating the controlled attenuation parameter (CAP) from ultrasound-guided attenuation parameter (UGAP)-attenuation coefficient (AC) and to evaluate the agreement and interchangeability between the two measurements in a clinical setting.

**Methods:** This retrospective observational study included 85 patients who underwent both UGAP and CAP measurements. UGAP-AC values were converted to dB/m to match CAP units. Pearson’s correlation and linear regression analyses were performed to derive a prediction formula. Bland–Altman analysis and repeated-measures analysis of variance were used to assess fixed and proportional biases and the influence of body mass index (BMI).

**Results:** UGAP-AC and CAP showed a strong correlation (*r* = 0.947, *p* < 0.0001). The regression equation was: CAP (dB/m) = 371.5 × UGAP-AC (dB/cm/MHz) + 12.3 (*R*^2^ = 0.8946). Bland–Altman analysis revealed a fixed bias of 28.6 dB/m and a BMI-dependent proportional bias. However, no significant proportional bias was observed within the clinically relevant CAP range (228–300 dB/m).

**Conclusion:** UGAP-AC can reliably estimate CAP using a simple regression formula. Although systematic biases exist, their clinical impact appears limited, supporting the interchangeable use of UGAP and CAP in routine practice.

## 1 Introduction

Metabolic dysfunction-associated steatotic liver disease (MASLD), formerly known as non-alcoholic fatty liver disease, is a chronic liver disease affecting approximately 1.7 billion people worldwide [1,2]. Among patients with MASLD, 20–30% may develop metabolic dysfunction-associated steatohepatitis (MASH), an inflammatory subtype that can lead to progressive fibrosis and ultimately cirrhosis [3]. Therefore, accurate identification of steatotic liver disease (SLD) is a critical initial step in clinical management.

In daily clinical practice, hepatic steatosis is most commonly assessed by B-mode ultrasound grading [4]. However, this method is subjective and lacks reproducibility. To overcome these limitations, ultrasound-based attenuation techniques have been developed to quantitatively evaluate fat accumulation by measuring the attenuation of ultrasound beams through the liver parenchyma [5]. The controlled attenuation parameter (CAP), which uses vibration-controlled transient elastography (VCTE; FibroScan, Echosens, Paris, France), was the first widely adopted noninvasive method and is considered a promising diagnostic tool for hepatic steatosis [6,7]. More recently, two-dimensional attenuation imaging techniques, such as ultrasound-guided attenuation parameter (UGAP; GE Healthcare, Milwaukee, USA), have been introduced as integrated applications in conventional ultrasound systems [8].

Given its widespread use and accumulating evidence, CAP is often regarded as the gold standard for ultrasound-based hepatic fat quantification. However, FibroScan functions as a dedicated and costly device, limiting its availability to advanced medical centers [9].

Additionally, performing serial evaluations using the same equipment often proves impractical in real-world settings due to device malfunctions or increased testing demand. Therefore, establishing the relationship between attenuation coefficients (AC) obtained by UGAP and CAP would support interchangeable use across platforms.

To date, no study has specifically addressed the compatibility and bias between these two widely used attenuation measurements. This study aimed to clarify the relationship between UGAP-AC and CAP and to derive a regression equation to estimate CAP from UGAP-AC.

## 2 Patients and methods

### 2.1 Study design and participant recruitment

This cross-sectional study enrolled 159 patients who underwent UGAP evaluation between January 2022 and December 2023 at Fukuoka Tokushukai Hospital (Fukuoka, Japan). Among these, UGAP was performed in 156 patients with clinically suspected SLD. Three patients were excluded due to an inability to hold their breath (n=2) or irregular fatty infiltration (n=1), resulting in a final study population of 156 patients.

First, UGAP-AC values were compared with B-mode steatosis grading in all 156 patients. Next, a subgroup of 85 patients who also underwent CAP measurement on the same day was analyzed to compare UGAP-AC and CAP. A flowchart of the patient selection process is shown in Figure 1. The study protocol was reviewed and approved by the Institutional Review Board of Fukuoka Tokushukai Hospital (Approval Number, 240401), and informed consent was obtained from all patients.

**Figure 1.**
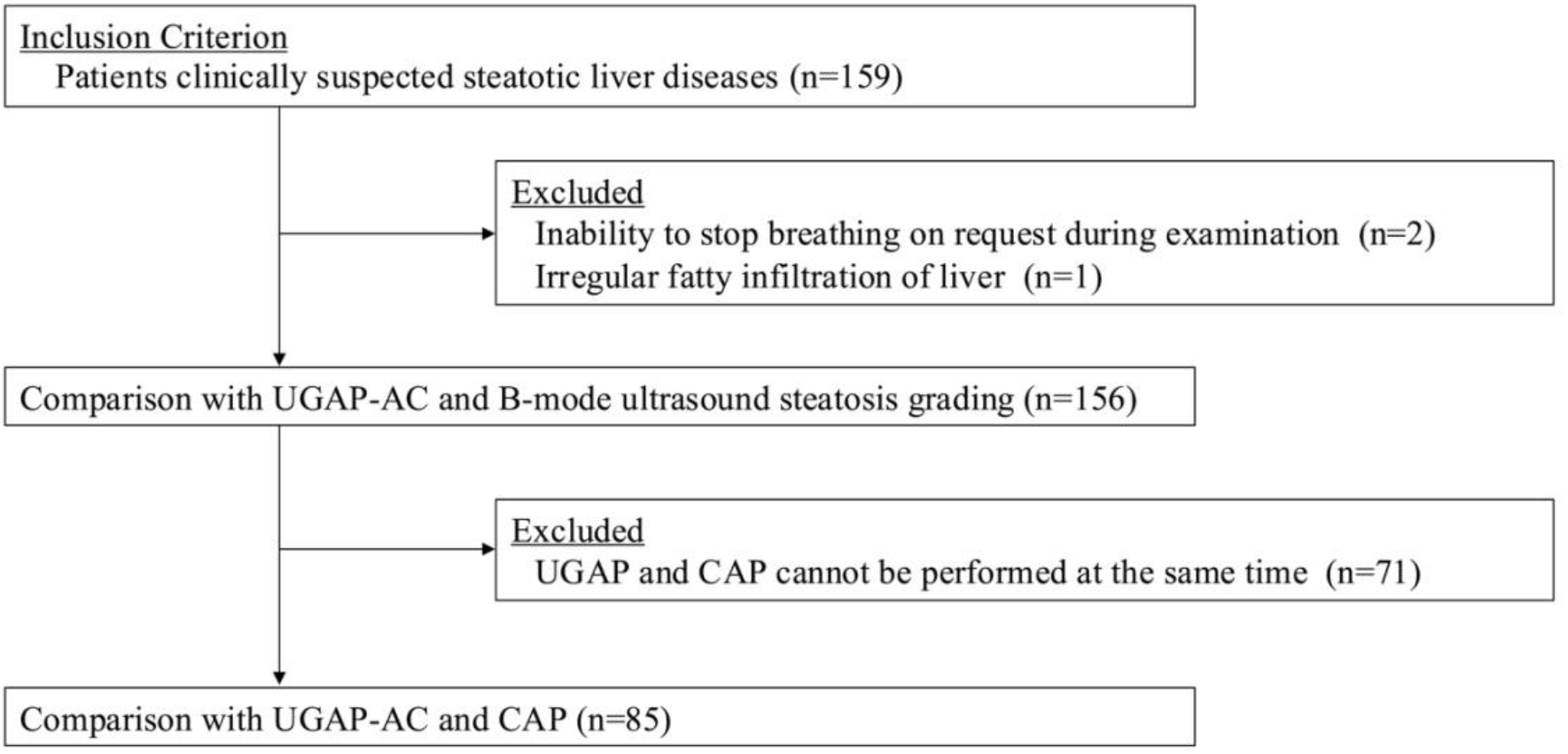
Flow diagram of patient enrollment. Abbreviations: AC, attenuation coefficient; UGAP, ultrasound-guided attenuation parameter; CAP, controlled attenuation parameter.

### 2.2 B-mode ultrasound steatosis grading

Hepatic steatosis was semi-quantitatively graded using a conventional B-mode assessment method [4] based on hepatorenal contrast, vascular blurring, and deep attenuation. Steatosis was categorized as mild, clear hepatorenal contrast; moderate, hepatorenal contrast with either vascular blurring or deep attenuation; and severe, hepatorenal contrast with both vascular blurring and deep attenuation.

### 2.3 UGAP measurement

UGAP was performed using the LOQIQ E10 ultrasound system (GE Healthcare, Milwaukee, WI, USA) equipped with a C1-6 transducer. All measurements were conducted by the first author, a sonographer with 15 years of experience in ultrasonography and formal training in UGAP.

The region of interest was placed in a homogeneous area of the right hepatic lobe, avoiding large vessels, and the “quality map” function was used to optimize image selection. A reliable UGAP measurement was defined as the median of 10 consecutive measurements obtained from the same region.

### 2.4 CAP measurement

CAP was measured using the Logiq S8+FS (GE Healthcare, Milwaukee, WI, USA) with integrated FibroScan. CAP measurement followed UGAP assessment and was performed at the same site by aligning the FibroScan M/XL probe under B-mode ultrasound guidance. A CAP interquartile range (IQR)/median ratio of < 0.3 was applied as a quality criterion for reliable measurements [10].

### 2.5 Statistical analyses

Numerical data were expressed as medians and IQRs, and categorical data were presented as numbers and percentages (n). Differences in UGAP-AC across B-mode steatosis grades were evaluated using the Kruskal–Wallis test. Normality was assessed using the Shapiro–Wilk test.

Pearson’s correlation coefficient was used to evaluate the association between the UGAP-AC and CAP. Bland–Altman analysis was performed to assess the agreement between the UGAP and CAP. Repeated-measures analysis of variance (ANOVA) was used to evaluate the interaction between the body mass index (BMI) group and attenuation measurements (UGAP vs. CAP).

## 3 Results

### 3.1 Patient population

The demographic characteristics of the participants are summarized in Table 1. Twenty-six patients who did not meet the ultrasound-based diagnostic criteria for hepatic steatosis were classified into the control group. Compared to the control group, the hepatic steatosis group showed significant differences in age, BMI, CAP, UGAP-AC, aspartate aminotransferase (AST), and alanine aminotransferase (ALT) (all *p* < 0.05). No significant differences were observed in platelet counts (PLT), and no evidence of fibrosis progression was found in the hepatic steatosis group.

**Table 1.** Demographic characteristics of enrolled patients Significant differences between the control and hepatic steatosis groups were determined using the Mann–Whitney U test for continuous variables and Fisher’s exact test for categorical variables. Abbreviations: AC, attenuation coefficient; ALT, alanine aminotransferase; AST, aspartate aminotransferase; T-Bil, total bilirubin; BMI, body mass index; CAP, controlled attenuation parameter; PLT, platelet count; UGAP, ultrasound-guided attenuation parameter.

| Variable |  | ALL | Control | Hepatic steatosis | p |
| --- | --- | --- | --- | --- | --- |
| Age (year) | n | 156 | 26 | 130 | P = 0.0181 |
|  | Median | 60 | 64 | 58 |  |
|  | Q1–Q3 | 48–70 | 60–73 | 44–69 |  |
| Sex | Male | 99 (63.5) | 15 (57.7) | 84 (64.6) | P = 0.5110 |
|  | Female | 57 (36.5) | 11 (42.3) | 46 (35.4) |  |
| BMI (kg/m<sup>2</sup>) | n | 146 | 26 | 120 | P = 0.0007 |
|  | Median | 25.1 | 23.7 | 25.7 |  |
|  | Q1–Q3 | 22.9–27.5 | 21.9–24.7 | 23.3–28.3 |  |
| CAP (dB/m) | n | 85 | 15 | 70 | P < 0.0001 |
|  | Median | 290 | 218 | 308 |  |
|  | Q1–Q3 | 266–326 | 207–228 | 278–333 |  |
| UGAP-AC (dB/cm/MHz) | n | 156 | 26 | 130 | P < 0.0001 |
|  | Median | 0.73 | 0.58 | 0.78 |  |
|  | Q1–Q3 | 0.67–0.85 | 0.56–0.60 | 0.70–0.86 |  |
| AST (U/L) | n | 151 | 26 | 125 | P = 0.0134 |
|  | Median | 28 | 23 | 29 |  |
|  | Q1–Q3 | 21–37 | 19–30 | 22–41 |  |
| ALT (U/L) | n | 149 | 26 | 123 | P < 0.0001 |
|  | Median | 32 | 22 | 37 |  |
|  | Q1–Q3 | 21–50 | 17–28 | 23–55 |  |
| T-Bil (g/dL) | n | 148 | 26 | 122 | P = 0.5770 |
|  | Median | 0.7 | 0.7 | 0.7 |  |
|  | Q1–Q3 | 0.6–1.0 | 0.5–1.0 | 0.6–1.0 |  |
| PLT (×10<sup>4</sup>/mm<sup>3</sup>) | n | 151 | 26 | 125 | P = 0.9590 |
|  | Median | 242 | 246 | 241 |  |
|  | Q1–Q3 | 200–277 | 197–283 | 201–271 |  |

### 3.2 Relationship between UGAP-determined attenuation coefficient and B-mode grading

The relationship between the UGAP-AC and B-mode steatosis grading is shown in Figure 2. The median UGAP-AC values for the control, mild, moderate, and severe B-mode grades were 0.58, 0.68, 0.71, and 0.85 dB/cm/MHz, respectively. Differences in UGAP-AC across the five B-mode steatosis grades were statistically significant (Kruskal–Wallis test: *p* < 0.0001), and post hoc pairwise comparisons revealed significant differences between all groups (Steel– Dwass test: all *p* < 0.0001).

**Figure 2.**
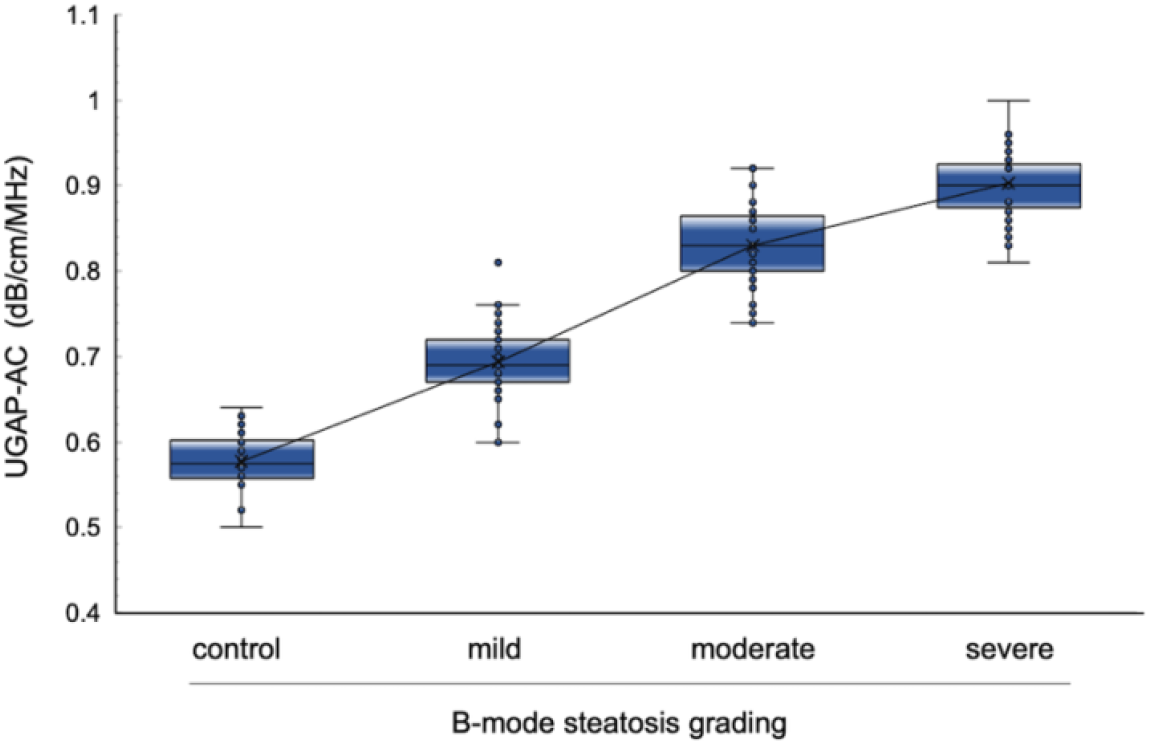
Relationship between UGAP-determined AC and B-mode staging. Box-and-whisker plots illustrate UGAP values across the B-mode-based hepatic steatosis grades (0–3). Boxes represent the interquartile range (IQR), horizontal lines indicate the median, and the “×” marks the mean. Whiskers extend to 1.5×IQR, with individual data points overlaid. The line connecting means shows a progressive increase across grades. UGAP-AC differed significantly among steatosis grades (Kruskal–Wallis test, *p* < 0.0001), with all pairwise comparisons also significant (Steel–Dwass test, all *p* < 0.0001). Abbreviations: AC, attenuation coefficient; UGAP, ultrasound-guided attenuation parameter.

### 3.3 Prediction of CAP using UGAP-AC

The Shapiro–Wilk test confirmed normal distributions for both CAP (*p* = 0.2885) and UGAP-AC (*p* = 0.0814). Pearson’s correlation analysis demonstrated a strong positive correlation between the two ACs (*r* = 0.947, 95% confidence interval [CI] = 0.919–0.965, *p* < 0.0001). As shown in Figure 3, a linear regression model was constructed with UGAP-AC (dB/cm/MHz) as the explanatory variable and CAP (dB/m) as the response variable, yielding the following equation: CAP = 371.5 × UGAP-AC + 12.3, *R*^*2*^ = 0.8946.

**Figure 3.**
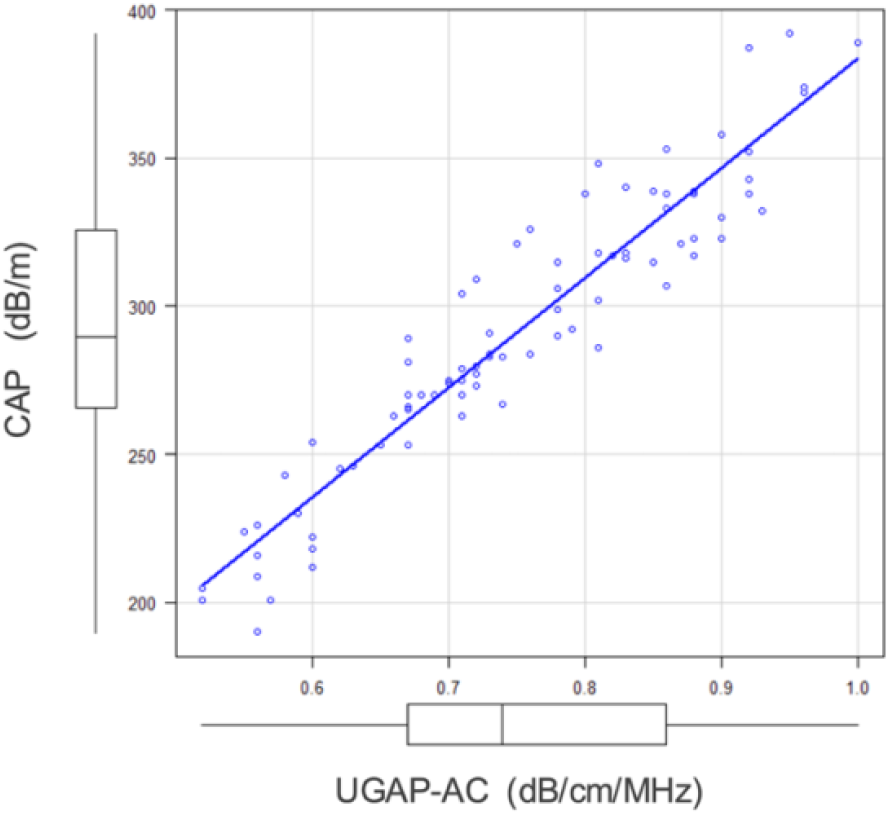
Correlation between UGAP-AC and CAP. Scatter plot demonstrating the strong positive linear correlation between the UGAP-AC and CAP (Pearson’s correlation coefficient r = 0.947, 95% CI: 0.919–0.965, *p* < 0.0001). Box plots along each axis show the distribution of respective variables. Abbreviations: AC, attenuation coefficient; CAP, controlled attenuation parameter; UGAP, ultrasound-guided attenuation parameter; CI, confidence interval.

A logarithmic transformation of UGAP-AC was also explored due to its borderline normality (*p* = 0.0814), although no meaningful improvement in model fit was observed (*R*^2^ = 0.8966 vs. 0.8946). Therefore, the original linear model was retained for its superior clinical interpretability.

In a multivariable analysis including UGAP-AC, BMI, ALT, AST, total bilirubin, and PLT, only UGAP-AC and BMI remained significant predictors of CAP (*R*^*2*^ = 0.9055). UGAP-AC accounted for the majority of the variance (*p* < 0.0001), and BMI also showed a statistically significant association (*p* = 0.0029) (Table 2).

**Table 2.** Multiple linear regression analysis of predictors of CAP. Abbreviations: AC, attenuation coefficient; ALT, alanine aminotransferase; AST, aspartate aminotransferase; BMI, body mass index; CAP, controlled attenuation parameter; PLT, platelet count; T-Bil, total bilirubin; UGAP, ultrasound-guided attenuation parameter; CI, confidence interval.

| Variables | Estimate | Lower 95% CI | Upper 95% CI | Std. err. | t statistic | p value |
| --- | --- | --- | --- | --- | --- | --- |
| (Intercept) | -10.352 | -40.868 | 20.164 | 15.325 | -0.675 | 0.5014 |
| AST | 0.037 | -0.117 | 0.190 | 0.077 | 0.475 | 0.6364 |
| ALT | -0.014 | -0.120 | 0.091 | 0.053 | -0.267 | 0.7899 |
| T-Bil | -2.170 | -11.031 | 6.691 | 4.450 | -0.488 | 0.6272 |
| PLT | -0.045 | -0.105 | 0.014 | 0.030 | -1.509 | 0.1354 |
| BMI | 1.651 | 0.584 | 2.719 | 0.536 | 3.082 | 0.0029 |
| UGAP-AC | 361.254 | 330.505 | 392.004 | 15.442 | 23.394 | < 0.0001 |

Given the balance between model accuracy and clinical applicability, the univariate regression model using UGAP-AC alone was adopted to construct the conversion equation.

### 3.4 Agreement and comparability of CAP and UGAP values

To enable direct comparison, UGAP-AC was converted from dB/cm/MHz to dB/m to match the units of CAP. The mean UGAP value was 264 ± 42 dB/m, and the mean CAP value was 292 ± 47 dB/m.

Agreement between the two attenuation methods was assessed using Bland–Altman analysis. The mean difference (bias) was 28.6 dB/m (95% CI: 25.3–31.9), with upper and lower limits of agreement (LOA) of 58.8 dB/m (95% CI: 54.0–66.0) and -1.5 dB/m (95% CI: -8.7–3.3), respectively. The precision was 15.7 dB/m. The Bland–Altman plots showed an approximately normal distribution of differences, with 95% of data points falling within the LOA, indicating good agreement between the two measurements (Figure 4A). A significant fixed bias was observed (*p* < 0.0001; 95% CI: 25.3–31.9 dB/m).

**Figure 4.**
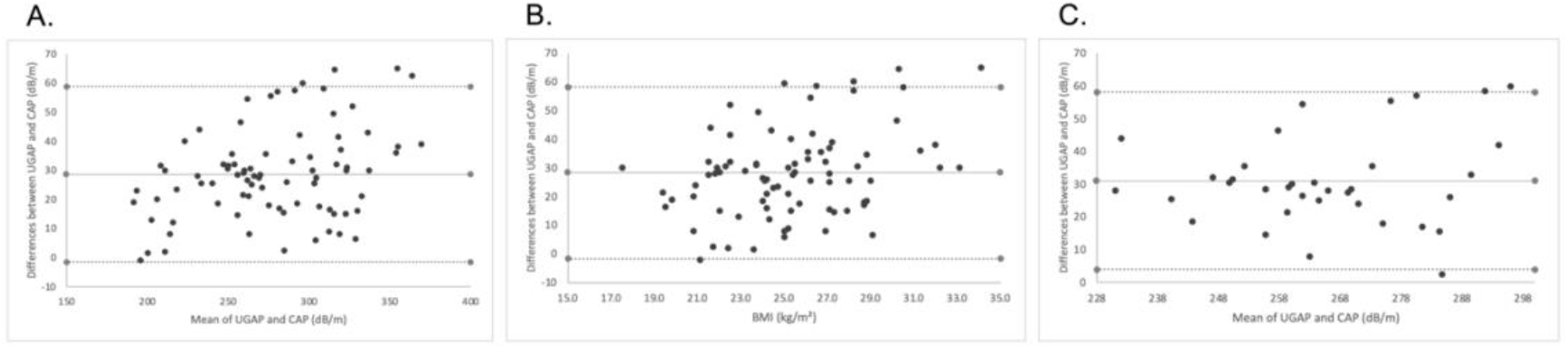
Bland–Altman analysis of agreement between UGAP and CAP measurements. Bland–Altman plot comparing UGAP and CAP measurements. The mean difference (bias) was 28.6 dB/m, with 95% limits of agreement ranging from −1.5 to 58.8 dB/m. The difference between UGAP and CAP was plotted against BMI to assess proportional bias. A significant positive association was observed between BMI and the difference (*p* = 0.0015). When the analysis was restricted to the clinically relevant CAP range (228–300 dB/m), the proportional bias was no longer significant (*p* = 0.2929), suggesting negligible bias in the diagnostic range. Abbreviations: BMI, body mass index; CAP, controlled attenuation parameter; UGAP, ultrasound-guided attenuation parameter.

Proportional bias was also identified, as evidenced by a significant correlation between the mean of the two methods and their differences (*r* = 0.12, *p* = 0.0018). To further investigate this bias, the x-axis was replaced with BMI, and the difference between UGAP and CAP was regressed against BMI. This analysis revealed a statistically significant proportional bias (*p* = 0.0015) (Figure 4B). However, when the analysis was restricted to the clinically relevant CAP range (228–300 dB/m), the proportional bias was no longer statistically significant (*p* = 0.2929), suggesting negligible bias in routine diagnostic use (Figure 4C).

### 3.5 Involvement of BMI in proportional bias

Given the proportional bias observed in the Bland–Altman analysis, the potential involvement of BMI was further examined. Participants were divided into two groups based on the mean BMI (< 25.3 or ≥ 25.3 kg/m^2^), and attenuation values from both UGAP and CAP were compared using repeated measures ANOVA.

The repeated measures ANOVA, with measurement method (UGAP vs. CAP) and BMI group as factors, revealed significant main effects of measurement method (*F*(1, 81) = 308.5, *p* < 0.0001) and BMI group (*F*(1, 81) = 4.61, *p* = 0.0348), along with a significant interaction between the method and BMI group (*F*(1, 81) = 7.93, *p* = 0.0061) (Figure 5).

**Figure 5.**
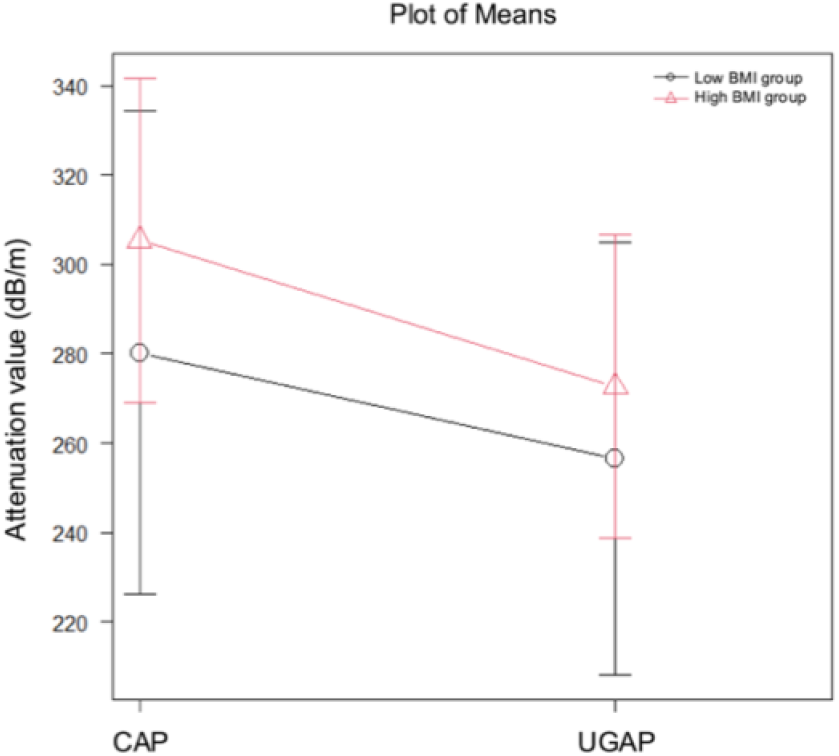
Interaction effect of BMI on attenuation values measured by CAP and UGAP. Mean attenuation values (± standard error) were measured by CAP and UGAP in two BMI groups: low BMI (< 25.3 kg/m^2^, black circles) and high BMI (≥ 25.3 kg/m^2^, red triangles). Repeated-measures ANOVA revealed significant main effects of the measurement method and BMI group, as well as a significant interaction between the two (p = 0.0061), indicating that the difference between CAP and UGAP values varied depending on BMI. Abbreviations: BMI, body mass index; CAP, controlled attenuation parameter; UGAP, ultrasound-guided attenuation parameter; ANOVA, analysis of variance.

## 4 Discussion

This study demonstrated a strong correlation and good agreement between UGAP and CAP measurements. Additionally, both fixed and proportional biases were identified, with the latter associated with BMI.

Hepatic steatosis refers to the accumulation of fat in ≥ 5% of hepatocytes [11].

Ultrasonography provides a simple and accessible modality for diagnosis; however, the sensitivity of B-mode imaging in detecting mild steatosis remains relatively low, estimated at approximately 61–65% [4,12,13]. Therefore, reliable screening for early-stage hepatic fat accumulation often requires quantitative ultrasound attenuation techniques [14].

Among these techniques, UGAP has been validated against liver biopsy [15-17] and MRI-PDFF [8] for its capacity to quantify hepatic fat content. However, in high-throughput community hospitals such as ours—where multiple ultrasound systems from various vendors are routinely used—standardizing the attenuation method across all patients often proves impractical. CAP, although considered reliable and widely accepted, involves higher costs and limited availability in such clinical environments. Establishing a conversion model between CAP and UGAP-AC would allow interchangeable use of these modalities; thereby, enhancing clinical utility in routine practice.

In this study, a linear regression model was developed to estimate CAP values from UGAP-AC measurements, aiming to enable the interchangeable use of these two attenuation parameters. The resulting equation demonstrated a strong linear relationship (*R*^2^ = 0.8946), supporting the feasibility of UGAP-AC as a surrogate marker for CAP. A previous study reported a correlation of *r* = 0.73 between UGAP and CAP [18], which did not suffice to justify compatibility or support the creation of a conversion model. In contrast, the much stronger correlation observed in this study (*r* = 0.947) provided a robust foundation for deriving an interchangeable equation between the two methods.

Although a log-transformed model was also explored due to the borderline normality of UGAP-AC, the improvement in model fit was negligible (*R*^2^ = 0.8966). Therefore, the original linear model was retained for its clinical simplicity and interpretability.

Furthermore, a multivariable analysis was performed to assess potential confounding factors such as BMI and liver enzyme levels. Although BMI emerged as a statistically significant covariate, its inclusion led to only a modest increase in explanatory power (*R*^2^ = 0.9055). Given the limited benefit and increased complexity, the univariable model using UGAP-AC alone was adopted. This approach enhances clinical applicability by enabling real-time CAP estimation based solely on routinely acquired UGAP measurements without requiring additional patient data.

Bland–Altman analysis identified both fixed and proportional biases, indicating systematic differences between UGAP and CAP measurements. The fixed bias likely stems from differences in calibration methods and the accuracy of acoustic property correction. UGAP relies on phantom-based calibration, whereas the CAP system does not disclose any equivalent correction process. The fixed bias identified in this study is reflected in the intercept of the regression equation, providing a basis for adjusted comparability between the two attenuation parameters.

To further investigate the proportional bias, the Bland–Altman plot was reanalyzed using BMI as the horizontal axis. A statistically significant but weak correlation emerged between BMI and the measurement differences (*r* = 0.12, *p* = 0.0018), suggesting that BMI may influence proportional bias, although the practical impact appears limited. Repeated measures ANOVA stratified by BMI group confirmed a significant interaction, with larger differences between CAP and UGAP values observed in patients with higher BMI.

This discrepancy may be attributed to technical differences in the measurement principles: UGAP automatically adjusts measurement depth along the centerline, whereas CAP uses a fixed depth. In patients with obesity, CAP may be more affected by subcapsular artifacts. Importantly, when the analysis was restricted to the clinically relevant CAP range of 228–300 dB/m—which includes diagnostic thresholds such as 248 dB/m for S1 and 280 dB/m for S3 [19]—the proportional bias no longer reached statistical significance. This finding suggests that UGAP and CAP remain compatible within the clinically important range. To our knowledge, this type of bias between the two attenuation methods has not been previously reported and represents a novel contribution of this study.

Although the regression model showed excellent performance in this study, external validation remains necessary. This study was conducted at a single institution using a specific ultrasound platform, underscoring the need for additional evaluation across other clinical settings and ultrasound systems.

The following regression equation was established to estimate CAP from UGAP-AC: CAP (dB/m) = 371.5 × UGAP-AC (dB/cm/MHz) + 12.3.

Although fixed and BMI-dependent proportional biases were identified between the two measurements, the proportional bias did not reach statistical significance within the clinically relevant CAP range. These findings support the interchangeable use of UGAP and CAP in routine clinical practice, particularly in settings where CAP remains unavailable.

## Data Availability

All data produced in the present study are available upon reasonable request to the authors

## Acknowledgments

The author thanks the technical support team at GE Healthcare for their valuable input on acoustic correction and measurement variability.

